# Early pre-exposure prophylaxis (PrEP) discontinuation among pregnant and postpartum women: Implications for maternal PrEP roll out in South Africa

**DOI:** 10.1101/2021.05.04.21256514

**Authors:** Dvora Leah Joseph Davey, Rufaro Mvududu, Nyiko Mashele, Maia Lesosky, Nehaa Khadka, Jessica More, Linda-Gail Bekker, Pamina Gorbach, Thomas J. Coates, Landon Myer, on behalf of PrEP-PP study team

## Abstract

**Introduction:** Oral pre-exposure prophylaxis (PrEP) is a safe and effective prevention strategy to reduce women’s risk of HIV in pregnancy and postpartum. Effective PrEP requires daily PrEP adherence, but little is known about maternal PrEP continuation and risk factors that influence optimal PrEP use.

**Methods:** The PrEP in pregnancy and postpartum (PrEP-PP) study is an ongoing cohort study that enrolled consenting pregnant, HIV-uninfected women at first antenatal care (ANC) visit, followed through 12-months postpartum. HIV-uninfected women and girls ≥16-years who were eligible for the study received HIV prevention counseling and were offered PrEP. Interviewers collected socio-demographic, behavioral data from participants at each visit. We analyzed the proportion of women who initiated PrEP and the proportion who continued on PrEP after 3-months with associated correlates, including side effects whilst on PrEP, by estimating the prevalence ratio (95% CI) adjusting for *a priori* confounders.

**Results:** Between Aug’19 and Feb’21, we enrolled 891 pregnant women (median gestation=21wks; age=26yrs). Following PrEP counseling, 90% of women initiated PrEP at their first ANC visit (n=801); 60% were married or cohabiting. Three-quarters of women on PrEP returned for a repeat prescription at 1-month; 62% returned at 3-months. One-third of women on PrEP reported a side effect, mostly nausea/vomiting, dizziness, and headache. Women on PrEP in the 1^st^ or 2^nd^ trimester had higher odds of reporting side effects (aOR=2.61; 95%CI=1.17-5.84) vs. postpartum women. Women who reported side effects continued with PrEP less than those who did not report side effects (aPR=0.88; 95% CI=0.78-0.99) adjusting for covariates. Women who had ≥1 previous pregnancy (aPR=0.76;95%CI=0.59-1.00) or were postpartum (aPR=0.86;95%CI=0.75-0.99) continued less than women who were primigravid or pregnant. Women who reported having an HIV+ partner (aPR= 1.70;95% CI=1.55-1.86) or unknown partner serostatus (aPR=1.14;95%CI=1.01-1.29) were more likely to continue on PrEP than those who had HIV-negative partners.

**Conclusion:** PrEP initiation and early continuation were high in ANC in this setting. Being postpartum and experiencing side effects were associated with lower PrEP continuation, presenting an opportunity for improved clinical management and counseling during pregnancy of nausea/vomiting to address early, transient side effects. Interventions for postpartum women on PrEP are urgently needed.

**Clinical Trial Number:** NCT03826199

## Introduction

Women in sub-Saharan Africa face a high risk of HIV acquisition during pregnancy and the breastfeeding period.(1) Evidence suggests that HIV acquisition risk more than doubles for women during pregnancy and breastfeeding.(2) While the elimination of mother-to-child transmission (EMTCT) services have expanded rapidly in the region, few prevention interventions exist for the majority of pregnant women who initially test HIV-negative in antenatal care (ANC).(3, 4) This is a major missed opportunity that has implications for women, their partners, and infants. Among women living with HIV, acute maternal HIV infection during pregnancy and breastfeeding substantially increases the risk of vertical transmission.(5) According to recent mathematical modelling, South Africa expects over 76,000 infant HIV cases between 2020 and 2030.(6, 7) These models demonstrated that pre-exposure prophylaxis (PrEP) provision may reduce perinatal HIV by 41% if 80% of *all* HIV-uninfected pregnant women use PrEP in pregnancy and breastfeeding.(6)

PrEP is a promising intervention to prevent HIV acquisition, with benefits both to the individual and population-level health. The World Health Organization (WHO) recommends offering PrEP to pregnant and postpartum women at risk for HIV acquisition as an individual-controlled prevention strategy.(8-10) Currently, PrEP counselling and services for cis-gender women, including those who are pregnant or breastfeeding, remain limited. Outside sub-Saharan Africa, most PrEP programmes are focused on men who have sex with men (MSM). The United States (US), Kenya, and South Africa have the greatest use of PrEP by women. Although over 300,000 people globally initiated PrEP since 2012, only a minority of them were women. (11)

A recent systematic review found that there is no safety-related rationale for prohibiting PrEP during pregnancy and/or breastfeeding.(7) There is minimal penetration of tenofovir (the metabolite of tenofovir disoproxil fumarate [TDF]) into breastmilk.(12) While safety data are reassuring, more data on how best to provide and optimize PrEP use in pregnancy and the postpartum period are needed. Oral PrEP is a safe and effective prevention strategy to reduce women’s risk of HIV in pregnancy and postpartum. There are multiple large-scale PrEP in pregnancy programs ongoing in South Africa and Kenya.(7, 13, 14). Effective PrEP requires daily PrEP adherence, but little is known about how minor symptoms, which may be more common during pregnancy, overlap with PrEP side effects and could impact PrEP persistence.

PrEP adherence and continuation have been low in women in South Africa,(15-19) and interventions are needed to improve PrEP uptake and adherence, especially in women at risk of HIV acquisition during pregnancy and breastfeeding.(20) There are numerous barriers to optimal maternal PrEP use in women across health facility-, intrapersonal-, and interpersonal-levels of population health. Common side effects, including gastrointestinal side effects (i.e. nausea, vomiting, diarrhoea) or dizziness are common in the early stages of taking PrEP. In Kenya, a frequent reason for discontinuation of PrEP in pregnant women (13) and adolescent girls and young women (AGYW) were side effects and low HIV risk perception (21). A recent meta-analysis of PrEP scale-up research in sub-Saharan Africa demonstrated that side effects were a key influence on PrEP use. (22)

To offer women additional choices to daily oral PrEP, alternative modalities are being evaluated, including long-acting injectables and intra-vaginal rings. Data from the HPTN084 trial showed a clear protective benefit from cabotegravir long-acting PrEP, with an 89 percent risk reduction compared to oral PrEP. The HPTN 084 trial will follow women who conceive in the study to evaluate the safety of cabotegravir in pregnant and breastfeeding women to inform PrEP in pregnancy studies. However, interventions to ensure high adherence are essential in daily TDF/FTC, vaginal rings, and in 3-monthly doses because of the long drug tail that could increase drug resistance in the case of skipped doses.(23)

Our study evaluated early PrEP initiation and continuation in a maternal PrEP cohort in a busy antenatal clinic in Cape Town, South Africa. We evaluated correlates of PrEP initiation and continuation in a cohort of pregnant and breastfeeding women to inform the national rollout of maternal PrEP programs in South Africa. Specifically, we evaluated the hypothesis that common side effects in pregnancy among women on PrEP may be a barrier to optimal PrEP use in pregnant women in our PrEP in pregnancy cohort in South Africa. Raising awareness on PrEP side effects amongst pregnant women and their healthcare providers, minimizing gaps in side effect management, and ensuring adherence and persistence of PrEP during periods of risk are critical issues if PrEP can have a meaningful impact on reducing HIV incidence in pregnant and breastfeeding women. Planning for, proposing, and promoting optimal PrEP use necessitates a deeper understanding of user barriers, including side effects, to ensure successful interventions.

## Methods

The PrEP-PP (PrEP in Pregnant and Postpartum women) study is an ongoing open prospective cohort that enrolls consenting pregnant, HIV-uninfected adolescent girls and women (age >=16 years) at the first ANC visit and follows participants through 12-months post-delivery from one public health clinic in Cape Town, Western Cape, South Africa. Recruitment began in August 2019 and is ongoing (N=891 as of February 28, 2021), with a planned sample size of 1,200 women.

### Study participants

Study eligibility criteria include: 1) age of ≥16 years, 2) confirmed HIV-negative serostatus by a 4^th^ generation antigen/antibody combination HIV test, 3) confirmed pregnancy status, 4) intention to stay in Cape Town through the postpartum period, 5) absence of contraindications to PrEP. Health care providers at study facilities provide group counselling to all pregnant women at baseline, which includes information on HIV testing and counselling, antiretroviral therapy (ART) for EMTCT, and the importance of HIV prevention for women who are HIV-uninfected. Eligible, consenting participants received 120 Rand (approximately $8 USD) in grocery vouchers for their time and effort in the study, as well as remuneration for transportation costs.

### Data collection

Following South African HIV testing guidelines, HIV counsellors provided ANC attendees without previous HIV diagnosis with rapid HIV testing and post-test counselling. (25) Upon confirmation of HIV seronegative status, trained study staff approach women to introduce the HIV prevention study. Upon agreement to participate in the study, the participant consented to screen for study eligibility, which included a rapid HIV antigen/antibody test and an HBsAg test (Abbott Laboratories). Upon eligibility confirmation and unassisted study consent, participants completed the baseline visit survey, which took 30 to 45 minutes using REDCap, a secure, web-based database platform. (26,27) Participants also received individual counselling about HIV prevention in pregnancy, including PrEP, along with information on consistent and correct condom use, knowing her partner’s HIV status (including referral for a male partner or couple’s HIV testing and counselling), and risk of serodiscordance. At baseline, participants self-collected a vaginal swab that is tested for *Chlamydia trachomatis (CT), Neisseria gonorrhoeae (NG)*, and *Trichomonas vaginalis (TV)* using point of care testing (Cepheid, Inc., Sunnyvale, CA, US) and treatment was provided following National STI Guidelines. (28) Data presented is through November 2020 when we stopped point-of-care testing as Cepheid was unable to place the order for STI kits.

Following the baseline survey, the study interviewer provided information about what PrEP is and the benefits of taking PrEP. The interviewer then asked the participant if they were interested in starting PrEP and disclosed that hesitancy or disinterest in PrEP initiation would not impact study participation. For study participants who agreed to initiate PrEP, the study nurse drew blood to measure baseline creatinine levels, results for which are confirmed within 24-48 hours. The nurse provided the patient with a one-month supply of Truvada® (tenofovir disoproxil fumarate/emtricitabine [TDF-FTC]) and an invitation card to return in one month for follow up testing (after which participants received a three-month prescription to correspond with quarterly study follow-up visits). Participants who did not start PrEP received an invitation to return in three months for a quarterly study follow-up visit. Follow-up visits were every 3-months and coincided with ANC visits until birth and the first postpartum visit. Follow-up visits lasted approximately 20-30 minutes.

### Data collection

Survey measures were collected at baseline and follow-up visits for all participants (on PrEP and not on PrEP). Survey measures included questions on: (a) basic demographic information and obstetric history (baseline only), (b) partner HIV status, (c) sexual behaviours in the past month and past week (including the number of sex partners, type of sex, frequency of sex and condom use), (d) substance use from the Alcohol Use Disorders Identification Test (AUDIT) (31) and Drug Use Disorders Identification Test (DUDIT) (32), (e) HIV risk perception, (f) intimate partner violence (using the WHO IPV scale (33,34)), (g) perceived partner, community and social support for PrEP, and (h) for PrEP users only, questions related to PrEP adherence according to self-report (seven-day and 30-day recall) and pill count measures, side effects, adverse events, severe adverse events and birth outcomes (after participants have given birth) at follow-up visits.

We defined PrEP initiation as accepting the initial PrEP prescription at baseline (first ANC visit). We defined PrEP continuation as receiving a PrEP prescription at both baseline and the three-month follow-up visit.

### Analyses

As of February 28, 2021, we had recruited and enrolled 891 participants. We present distribution of PrEP initiation (PrEP prescription received or not at baseline), PrEP continuation at three months in those who initiated PrEP at baseline (returned to the three-month follow-up visit and received another PrEP prescription or missed the visit), including counts and percentages for categorical variables and median and interquartile range (IQR) for continuous variables.

We present demographic characteristics including age, education, and marital status. We also include gestational age in weeks at baseline, gravidity (number of prior pregnancies). We analyse sexual behaviour data, including partner HIV testing in the past year, partner HIV status, condom use at last sex, and number of sexual partners during pregnancy. Finally, we selected substance use characteristics including any drug or alcohol use in the last year before pregnancy using the AUDIT and DUDIT scales.

We assess potential confounders with directed acyclic graphs (DAGs) (Supplemental Material). Age, marital status, gestational age at baseline are included as *a priori* confounders in the multivariable analysis of PrEP initiation and continuation with the exposures and outcomes of interest (1. PrEP initiation at baseline, 2. PrEP continuation at three months, and 3. PrEP adherence at three months). We also present side effects reported in women on PrEP at 3 months and reasons for PrEP discontinuation by age (<24 years vs. =>24 years).

We construct univariate and multivariable poisson regression models to evaluate correlates of PrEP initiation in pregnant women at baseline and PrEP continuation at 3-months using a two-tailed test to evaluate significance, with a significance threshold of p<0.05. All statistical analyses were conducted with STATA v.15.

### Ethics

The PrEP-PP study was approved by the Human Research Ethics Committee at the University of Cape Town (#297/2018) and by the University of California, Los Angeles Institutional Review Board (IRB#18-001622).

## Results

### Cohort characteristics

Between August 2019 and February 2021, we enrolled 891 pregnant women at their first ANC visit. The median age was 26 years (IQR 22-30) and the median gestational age was 21 weeks (IQR=14-31). Over half of women in the cohort had completed secondary school or higher (n=459, 52%). Almost all had one or more prior pregnancy (n=866, 97%). Fifty-six percent of participants who had a partner were cohabiting or married (n=499) and 17% reported having more than 1 sex partner in the past 12 months (n=151). Most women reported that their partner was HIV-negative (66%) or they didn’t know their serostatus (33%). At baseline, 90% of women were sexually active during pregnancy (n=805), of which 30% reported using a condom at last sex (n=264). Overall, 13% of women reported experiencing emotional, physical or psychological intimate partner violence (IPV). Half reported alcohol and/or drug use in the past year prior to pregnancy (48%; n=430). One-third of women were diagnosed with one or more STIs including CT, NG, and/or TV. **(Table 1)**

**Table 1.**
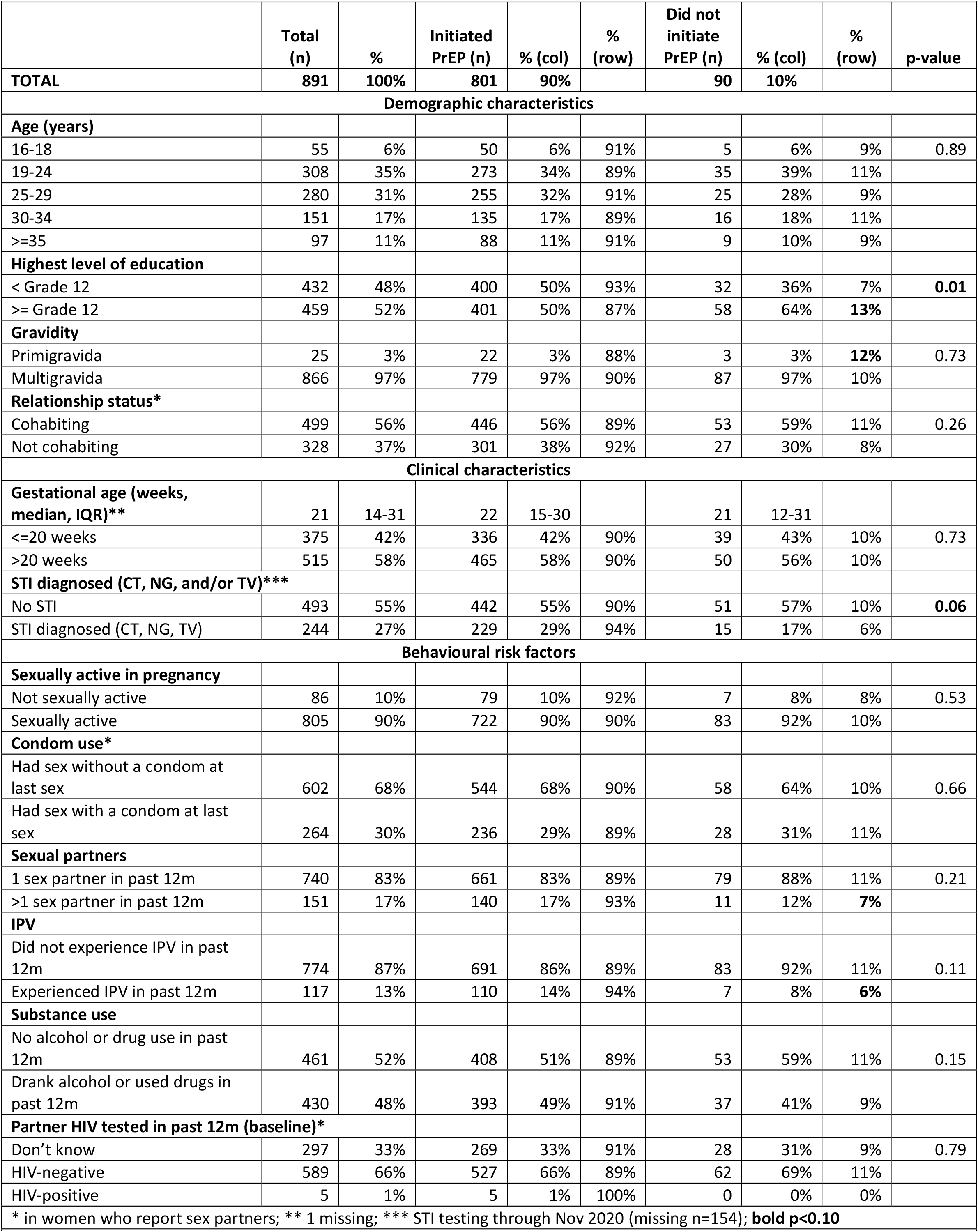
Characteristics of pregnant women offered PrEP in ANC in Cape Town, South Africa (N=891)

### PrEP initiation

Following PrEP counseling, 90% of women initiated PrEP at their first ANC visit (n=801), including n=50 young women 16-18 years old. Pregnant women who initiated PrEP at the first ANC visit had a lower level of education. Overall, 87% of women who had a secondary level of education initiated PrEP compared with 93% of women with a lower level education (p=0.01). Women diagnosed with a STI were more likely to initiate PrEP at initiation (94% vs 90%; p=0.058). Pregnant women who experienced IPV in the past year also initiated PrEP more than those who did not experience IPV (94% vs. 89%, p=0.11), though this was not statistically significant.

### PrEP continuation

Overall, 75% of women on PrEP at baseline returned for a repeat prescription at 1-month (n=668), and 62% returned and continued on PrEP at 3-months (n=393 of 635 eligible for 3-month visit). Women who continued on PrEP did not differ by age, education, relationship status, or baseline gestational age. Pregnant and postpartum women who continued on PrEP at 3 months tended to be higher risk in terms of experiencing IPV in the last year (74% continued vs. 60% who did not report recent IPV) and substance users (66% continued vs. 57% who did not report recent substance use; p=0.02). Women who continued were predominately pregnant vs. postpartum, did not experience side effects, and were earlier in their pregnancy when they came for their first ANC visit and started PrEP. **(Table 2)**

**Table 2.** Characteristics of pregnant women on PrEP who persisted on PrEP at 3-months in ANC in Cape Town, South Africa (n=635)

|  | Total<br>(n) | % | Continued<br>on PrEP at 3-months (n) | %<br>(col) | %<br>(row) | Discontinued<br>at PrEP at 3-months (n) | %<br>(col) | %<br>(row) | p-<br>value |
| --- | --- | --- | --- | --- | --- | --- | --- | --- | --- |
| **Total** | **635** | **100** | **393** |  | **62%** | **242** |  | **38%** |  |
| **Demographic characteristics** |  |  |  |  |  |  |  |  |  |
| **Age (years)** |  |  |  |  |  |  |  |  |  |
| 16-18 | 39 | 6% | 24 | 6% | 62% | 15 | 6% | 38% | 0.98 |
| 19-24 | 228 | 36% | 142 | 36% | 62% | 86 | 36% | 38% |  |
| 25-29 | 199 | 31% | 120 | 31% | 60% | 79 | 33% | 40% |  |
| 30-34 | 97 | 15% | 61 | 16% | 63% | 36 | 15% | 37% |  |
| >=35 | 72 | 11% | 46 | 12% | 64% | 26 | 11% | 36% |  |
| **Education** |  |  |  |  |  |  |  |  |  |
| < Grade 12 | 315 | 50% | 199 | 51% | 63% | 116 | 48% | 37% | 0.51 |
| >= Grade 12 | 320 | 50% | 194 | 49% | 61% | 126 | 52% | 39% |  |
| **Gravidity** |  |  |  |  |  |  |  |  |  |
| Primigravida | 15 | 2% | 12 | 3% | 80% | 3 | 1% | 20% | 0.18 |
| Multigravida | 620 | 98% | 381 | 97% | 61% | 239 | 99% | 39% |  |
| **Relationship status\*** |  |  |  |  |  |  |  |  |  |
| Cohabiting | 361 | 57% | 227 | 58% | 63% | 134 | 55% | 37% | 0.49 |
| Not cohabiting | 233 | 37% | 140 | 36% | 60% | 93 | 38% | 40% |  |
| **Clinical characteristics** |  |  |  |  |  |  |  |  |  |
| **Gestational age (weeks, median, IQR)** | 21 | 15-30 | 20 | 14-29 |  | 23 | 15-32 |  |  |
| <=20 weeks | 262 | 41% | 172 | 44% | 66% | 90 | 37% | 34% | 0.10 |
| >20 weeks | 373 | 59% | 221 | 56% | 59% | 152 | 63% | 41% |  |
| **STI diagnosed (CT, NG, and/or TV)** |  |  |  |  |  |  |  |  |  |
| No STI | 394 | 62% | 242 | 62% | 61% | 152 | 63% | 39% | 0.97 |
| STI diagnosed | 216 | 34% | 133 | 34% | 62% | 83 | 34% | 38% |  |
| **Side effects reported at last visit** |  |  |  |  |  |  |  |  |  |
| Did not experienced side effects | 231 | 36% | 181 | 46% | 78% | 50 | 21% | 22% | **0.03** |
| Experienced side effects | 210 | 33% | 145 | 37% | 69% | 65 | 27% | 31% |  |
| **Pregnancy status** |  |  |  |  |  |  |  |  |  |
| Pregnant | 411 | 65% | 267 | 68% | 65% | 144 | 60% | 35% | **0.03** |
| Postpartum (gave birth since last visit) | 224 | 35% | 126 | 32% | 56% | 98 | 40% | 44% |  |
| **Behavioural risk factors (baseline)** |  |  |  |  |  |  |  |  |  |
| **Sexually active in pregnancy** |  |  |  |  |  |  |  |  |  |
| Not sexually active | 64 | 10% | 39 | 10% | 61% | 25 | 10% | 39% | 0.87 |
| Sexually active | 571 | 90% | 354 | 90% | 62% | 217 | 90% | 38% |  |
| **Condom use at last sex\*** |  |  |  |  |  |  |  |  |  |
| Condomless sex | 425 | 67% | 264 | 67% | 62% | 161 | 67% | 38% | 0.69 |
| Used a condom | 192 | 30% | 116 | 30% | 60% | 76 | 31% | 40% |  |
| **Sexual partners** |  |  |  |  |  |  |  |  |  |
| 1 sex partner in past 12m | 520 | 82% | 320 | 81% | 62% | 200 | 83% | 38% | 0.70 |
| >1 sex partner in past 12m | 115 | 18% | 73 | 19% | 63% | 42 | 17% | 37% |  |
| **IPV** |  |  |  |  |  |  |  |  |  |
| Did not experience IPV in past 12m | 546 | 86% | 327 | 83% | 60% | 219 | 90% | 40% | **0.01** |
| Experienced IPV in past 12m | 89 | 14% | 66 | 17% | 74% | 23 | 10% | 26% |  |
| **Substance use** |  |  |  |  |  |  |  |  |  |
| No alcohol or drug use in past 12m | 322 | 51% | 185 | 47% | 57% | 137 | 57% | 43% | **0.02** |
| Drank alcohol or used drugs in past 12m | 313 | 49% | 208 | 53% | 66% | 105 | 43% | 34% |  |

| Partner HIV tested in past 12m (baseline)* |  |  |  |  |  |  |  |  |  |
| --- | --- | --- | --- | --- | --- | --- | --- | --- | --- |
| Don't know | 208 | 32% | 140 | 36% | 67% | 68 | 27% | 33% | 0.04 |
| HIV-negative | 423 | 67% | 249 | 63% | 59% | 174 | 72% | 41% |  |
| HIV-positive | 4 | 1% | 4 | 1% | 100% | 0 | 0% | 0 |  |

\* Of those who reported having a recent partner

Women who reported having a partner living with HIV were more likely to continue on PrEP (100% continued vs. 59% of those with HIV-negative partners (aPR= 1.70, 95% CI=1.55, 1.86), and 69% of those with partners of unknown serostatus (aPR= 1.14, 95% CI=1.01, 1.29). Reporting intimate partner violence or substance use in the past year were both associated with an increased likelihood of continuing on PrEP in pregnant and postpartum women (aPR for IPV=1.24, 95% CI=1.08, 1.44; aPR for substance use=1.16, 95% CI=1.02, 1.32). **(Table 3)**

**Table 3.**
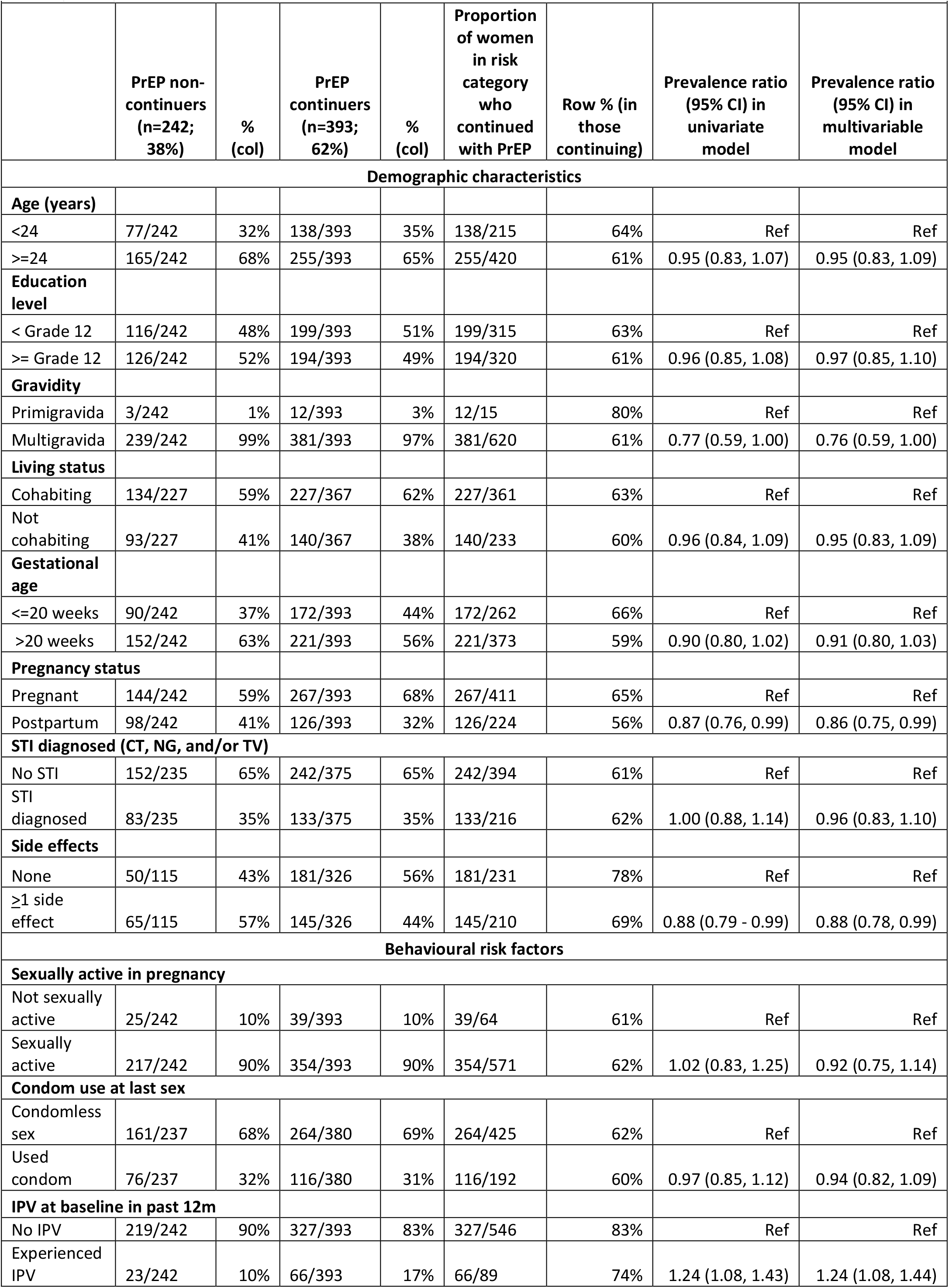
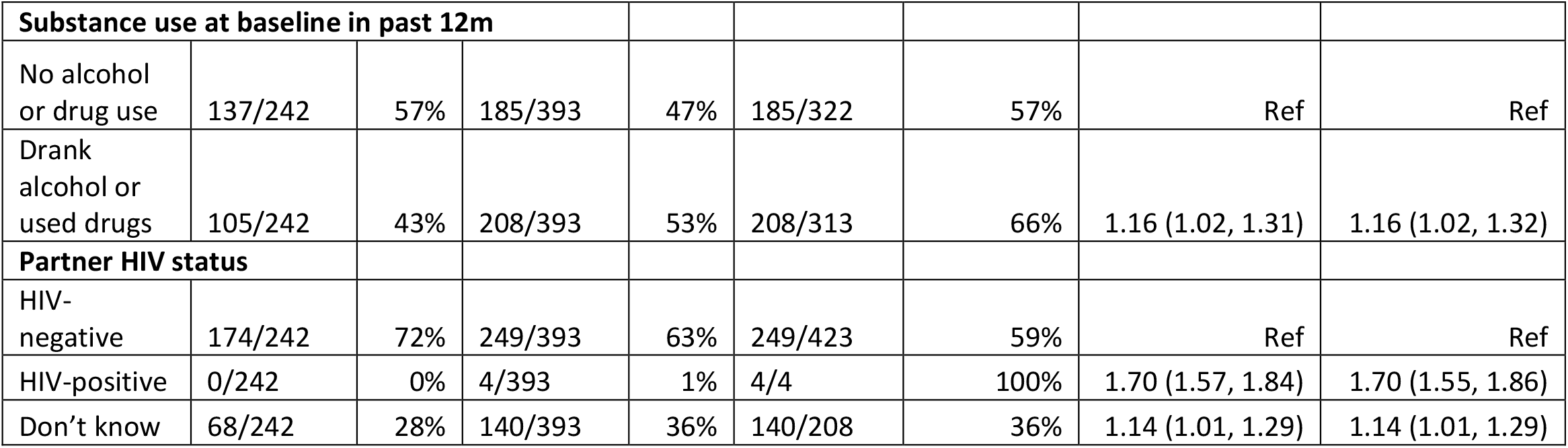
Correlates of PrEP continuation at 3 months among women who had initiated PrEP in ANC in Cape Town, South Africa (n=635)

|  | PrEP non-continuers<br>(n=242;<br>38%) | %<br>(col) | PrEP<br>continuers<br>(n=393;<br>62%) | %<br>(col) | Proportion<br>of women<br>in risk<br>category<br>who<br>continued<br>with PrEP | Row % (in<br>those<br>continuing) | Prevalence ratio<br>(95% CI) in<br>univariate<br>model | Prevalence ratio<br>(95% CI) in<br>multivariable<br>model |
| --- | --- | --- | --- | --- | --- | --- | --- | --- |
| **Demographic characteristics** |  |  |  |  |  |  |  |  |
| **Age (years)** |  |  |  |  |  |  |  |  |
| <24 | 77/242 | 32% | 138/393 | 35% | 138/215 | 64% | Ref | Ref |
| >=24 | 165/242 | 68% | 255/393 | 65% | 255/420 | 61% | 0.95 (0.83, 1.07) | 0.95 (0.83, 1.09) |
| **Education level** |  |  |  |  |  |  |  |  |
| < Grade 12 | 116/242 | 48% | 199/393 | 51% | 199/315 | 63% | Ref | Ref |
| >= Grade 12 | 126/242 | 52% | 194/393 | 49% | 194/320 | 61% | 0.96 (0.85, 1.08) | 0.97 (0.85, 1.10) |
| **Gravidity** |  |  |  |  |  |  |  |  |
| Primigravida | 3/242 | 1% | 12/393 | 3% | 12/15 | 80% | Ref | Ref |
| Multigravida | 239/242 | 99% | 381/393 | 97% | 381/620 | 61% | 0.77 (0.59, 1.00) | 0.76 (0.59, 1.00) |
| **Living status** |  |  |  |  |  |  |  |  |
| Cohabiting | 134/227 | 59% | 227/367 | 62% | 227/361 | 63% | Ref | Ref |
| Not cohabiting | 93/227 | 41% | 140/367 | 38% | 140/233 | 60% | 0.96 (0.84, 1.09) | 0.95 (0.83, 1.09) |
| **Gestational age** |  |  |  |  |  |  |  |  |
| <=20 weeks | 90/242 | 37% | 172/393 | 44% | 172/262 | 66% | Ref | Ref |
| >20 weeks | 152/242 | 63% | 221/393 | 56% | 221/373 | 59% | 0.90 (0.80, 1.02) | 0.91 (0.80, 1.03) |
| **Pregnancy status** |  |  |  |  |  |  |  |  |
| Pregnant | 144/242 | 59% | 267/393 | 68% | 267/411 | 65% | Ref | Ref |
| Postpartum | 98/242 | 41% | 126/393 | 32% | 126/224 | 56% | 0.87 (0.76, 0.99) | 0.86 (0.75, 0.99) |
| **STI diagnosed (CT, NG, and/or TV)** |  |  |  |  |  |  |  |  |
| No STI | 152/235 | 65% | 242/375 | 65% | 242/394 | 61% | Ref | Ref |
| STI diagnosed | 83/235 | 35% | 133/375 | 35% | 133/216 | 62% | 1.00 (0.88, 1.14) | 0.96 (0.83, 1.10) |
| **Side effects** |  |  |  |  |  |  |  |  |
| None | 50/115 | 43% | 181/326 | 56% | 181/231 | 78% | Ref | Ref |
| ≥1 side effect | 65/115 | 57% | 145/326 | 44% | 145/210 | 69% | 0.88 (0.79 - 0.99) | 0.88 (0.78, 0.99) |
| **Behavioural risk factors** |  |  |  |  |  |  |  |  |
| **Sexually active in pregnancy** |  |  |  |  |  |  |  |  |
| Not sexually active | 25/242 | 10% | 39/393 | 10% | 39/64 | 61% | Ref | Ref |
| Sexually active | 217/242 | 90% | 354/393 | 90% | 354/571 | 62% | 1.02 (0.83, 1.25) | 0.92 (0.75, 1.14) |
| **Condom use at last sex** |  |  |  |  |  |  |  |  |
| Condomless sex | 161/237 | 68% | 264/380 | 69% | 264/425 | 62% | Ref | Ref |
| Used condom | 76/237 | 32% | 116/380 | 31% | 116/192 | 60% | 0.97 (0.85, 1.12) | 0.94 (0.82, 1.09) |
| **IPV at baseline in past 12m** |  |  |  |  |  |  |  |  |
| No IPV | 219/242 | 90% | 327/393 | 83% | 327/546 | 83% | Ref | Ref |
| Experienced IPV | 23/242 | 10% | 66/393 | 17% | 66/89 | 74% | 1.24 (1.08, 1.43) | 1.24 (1.08, 1.44) |
| **Substance use at baseline in past 12m** |  |  |  |  |  |  |  |  |
| No alcohol or drug use | 137/242 | 57% | 185/393 | 47% | 185/322 | 57% | Ref | Ref |
| Drank alcohol or used drugs | 105/242 | 43% | 208/393 | 53% | 208/313 | 66% | 1.16 (1.02, 1.31) | 1.16 (1.02, 1.32) |
| **Partner HIV status** |  |  |  |  |  |  |  |  |
| HIV-negative | 174/242 | 72% | 249/393 | 63% | 249/423 | 59% | Ref | Ref |
| HIV-positive | 0/242 | 0% | 4/393 | 1% | 4/4 | 100% | 1.70 (1.57, 1.84) | 1.70 (1.55, 1.86) |
| Don't know | 68/242 | 28% | 140/393 | 36% | 140/208 | 36% | 1.14 (1.01, 1.29) | 1.14 (1.01, 1.29) |

### Side effects in pregnant women on PrEP

Overall, 210 women reported side effects at the 3-month follow up visit (48% of n=441 who attended the visit). The most common side effects were nausea and vomiting, reported by 39% and 26% of women who reported side effects, respectively. Approximately one in four women who reported nausea, vomiting, or a headache said that it bothered them “some” or “a lot”. While fewer women reported dizziness (16%), diarrhoea (2%), or bad dreams/insomnia (1%), these side effects bothered all women “some” or “a lot”. **(Figure 1)**. After one-month of follow-up, women in the 1st or 2nd trimester who were on PrEP had the highest odds of reporting side effects (aOR=2.61; 95%CI=1.17-5.84) compared to postpartum women, adjusting for maternal age (not tabled). One-third of women who reported a side effect continued on PrEP at 3-months, compared with 41% in those who did not experience side effects (aPR=0.88; 95% CI=0.79, 0.88) adjusting for age, gestational age at baseline, and relationship status. Women who had ≥1 prior pregnancy (aPR=0.76; 95% CI=0.59, 1.00) or were postpartum (aPR=0.86; 95% CI=0.75, 0.99) discontinued PrEP more than women who were primigravid or who were pregnant.

**Figure 1.**
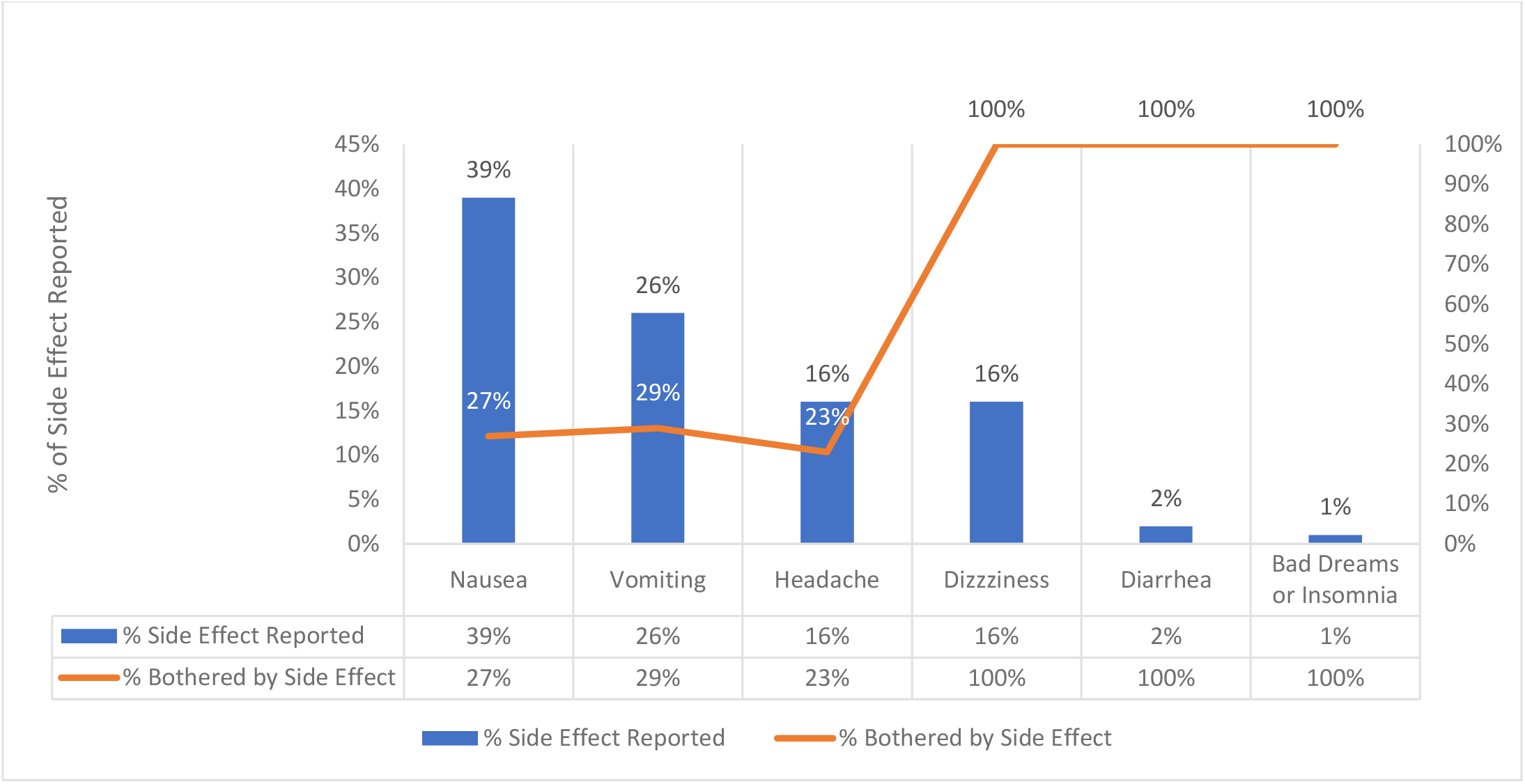
Side effects reported by pregnant women on PrEP and proportion reporting side effect bothered them

### Reason for missing PrEP doses

Women who reported taking PrEP but missing 1 or more doses were asked why they missed their PrEP doses in the past month. Out of 261 pregnant women, the most common reasons cited were because of forgetting (32%), due to travel (31%), or experiencing side effects and/or feeling ill (25%). About 1% of women indicated fear of safety and 11% of pregnant women who missed a dose of PrEP noted study or time burden. The only reason that was significantly associated with non-continuation of PrEP was side effects in the past month (p=0.04). None of the other reasons were associated with PrEP discontinuation. **(Figure 2)**

**Figure 2.**
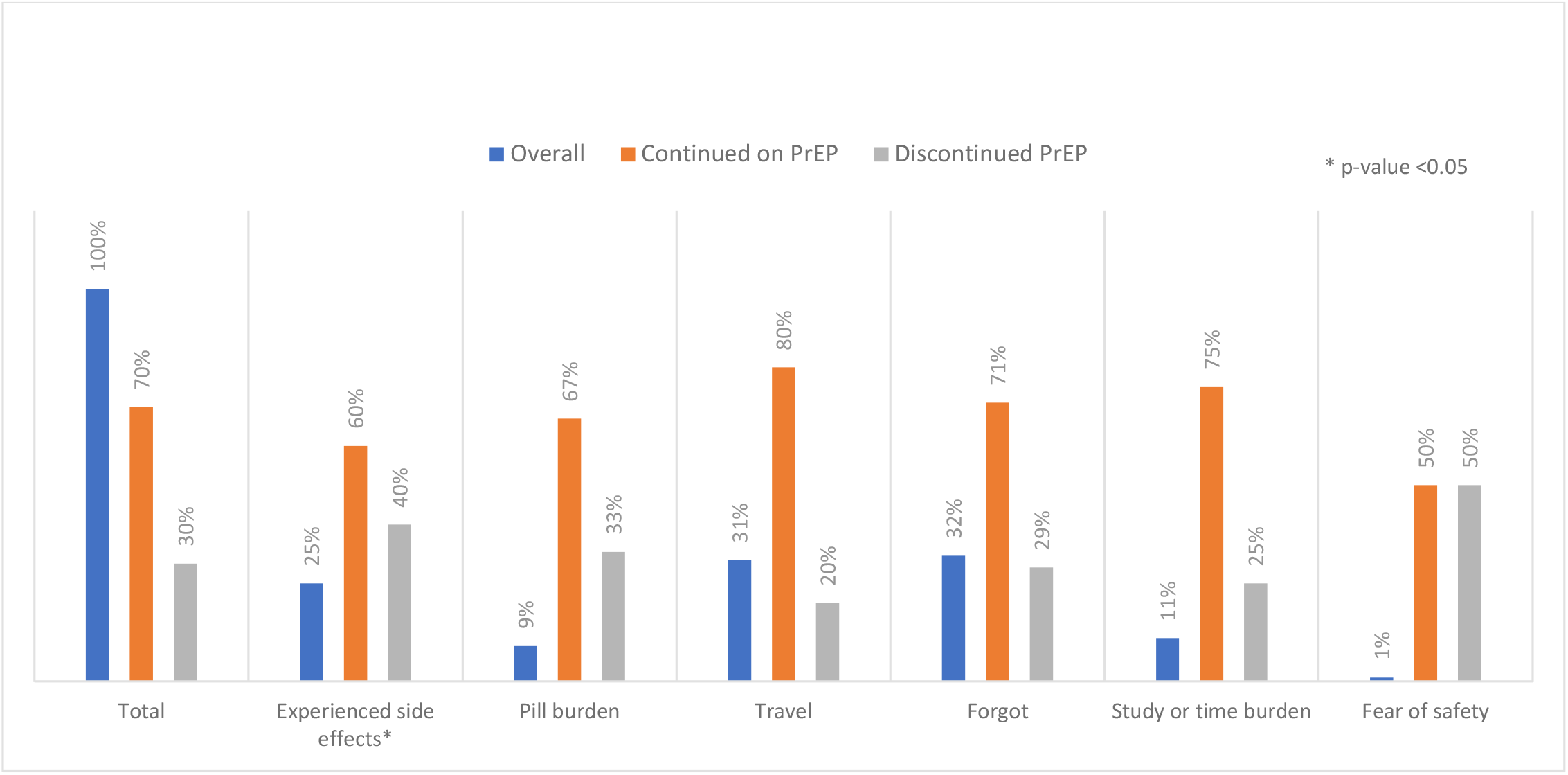
Reasons for missing PrEP doses among Pregnant and Postpartum women at 3 months

## Discussion

Our study is one of the first to integrate PrEP provision in ANC in a high HIV incidence community in South Africa. PrEP initiation was high in the ANC setting and especially among higher risk pregnant women, including those diagnosed with a STI at initiation. We identified important barriers to early PrEP continuation in pregnancy, including common side effects like nausea and vomiting that may be overlapping with pregnancy symptoms. In addition, we identified a significant drop off in PrEP continuation among postpartum women who no longer returned to the same clinic for their regular ANC visit. These results present opportunities for improved clinical management and counseling during pregnancy of nausea and vomiting to normalize early, transient side effects to improve PrEP adherence in pregnant women. Other opportunities include PrEP integration into maternal childcare and pediatric care to continue offering PrEP and counseling in breastfeeding women at risk of HIV acquisition.

PrEP was offered to HIV-uninfected pregnant women at their first antenatal visit (over 90% of those who were eligible and opted to be in the study). Almost 70% of women continued taking PrEP at 3-months after starting ANC. This proportion was significantly lower in postpartum women who were no longer attending regular ANC visits compared to pregnant women. Significant issues with PrEP continuation have been identified in other studies, with 50% or more of clients discontinuing within the first one to six months of use at sites in Kenya, South Africa, and the United States (24, 25). However, this proportion is higher than other populations of non-pregnant adolescent girls and young women (25) and female sex workers in South Africa (26), or pregnant women in lower HIV prevalence settings like in Kisumu, Kenya, where 39% of women continued on PrEP after one month and 41% in a family planning programme (13, 21). Similar proportions of PrEP continuation have been observed in African women in heterosexual serodiscordant couples (27), and in heterosexual women in the United States (28).

Our study identified important correlates of PrEP initiation and continuation in pregnancy. Importantly, we found that one-fifth of pregnant women reported side effects while taking PrEP and about half of those said that the side effects bothered them. Management of nausea and vomiting in pregnancy may be an important driver of PrEP continuation, including counseling women on the transient nature of side effects in pregnancy. Trained HIV counselors advised women to take PrEP at night, when they did not have morning sickness, or at a different time than other multi-vitamins and folic acid which may also cause nausea.

Having a male partner with an unknown HIV serostatus was associated with HIV acquisition in pregnancy and breastfeeding (29). Approximately 67% of women in our cohort knew their partner’s serostatus. Prior studies have shown that male partners are rarely engaged in pregnancy care or couples’ HIV testing in South Africa.(30) Innovative strategies to improve partner HIV testing may also optimize PrEP use in pregnant women by improving women’s awareness of her partner’s HIV status. In a Kenyan study, 53% of women reported that her partner used an HIV self-test, which may improve risk assessment and PrEP use.(31) PrEP programmes should consider how best to integrate HIV self-testing for monitoring of HIV status in PrEP users, and to offer couples HIV testing and encourage mutual disclosure in pregnant and breastfeeding women.

The most common reason for pregnant women to discontinue taking PrEP was because of side effects and pill burden (e.g., forgetting or travelling) associated with daily PrEP intake. Side effects and stigma were also cited as the most common reason for other South African men who have sex with men and sex workers to discontinue taking PrEP.(24) Long-acting PrEP methods are currently being tested in pregnant and breastfeeding women. However, daily pills may be preferred by some women who want control over when they start or stop taking PrEP, or who do not want injectables like injection or vaginal ring. Therefore, addressing the common side effects in PrEP users is essential to improving adherence. This consideration is essential in pregnant women who require daily dosing for efficacy, given that non-adherence is less forgiving than in postpartum women.(32)

Our study had several strengths. Integrating PrEP into a busy government community health center allowed us to understand how best to offer PrEP to HIV-uninfected pregnant women, and what common barriers exist for short-term continuation. However, our study has limitations as well. Firstly, we used trained study staff to provide counseling and nurses to prescribe PrEP which limits the real-life implementation in the health facility Secondly, there was a large proportion of women who were lost to follow up after initiating PrEP. This study may have under-estimated factors like lack of stable transportation from women’s residence to a health clinic, intimate partner violence, and the impact of the COVID-19 lockdown orders as other reasons for discontinuation. Finally, our study evaluated women returning for PrEP prescription as a proxy for PrEP continuation but it is not a measure of PrEP adherence. Future studies will evaluate tenofovir diphosphate levels to understand levels of objective adherence in this cohort.

## Conclusions

In conclusion, we found a significant proportion of pregnant women were interested in starting PrEP at their first antenatal visit, especially among higher risk pregnant women. Continuation on PrEP at 3-months was also high when compared to other South African populations but it was lower in postpartum women who do not return to the same facility for ANC visits, and in women who report side effects early on that overlap with pregnancy symptoms. Our study findings present opportunities for interventions to optimize PrEP use in pregnant and breastfeeding women, including improved counseling and clinical management around side effects to normalize early, transient side effects, and decentralized services to pediatric care or community services for postpartum, breastfeeding women.

## Data Availability

Data is available upon request from study investigators. Please for more information

## Competing interests

We received the study drug (Truvada^®^) from Gilead (California, USA) and STI test kits from Cepheid (California, USA).

## Authors’ contributions

DJD designed the study, analyzed the data, wrote the first draft of the paper, and reviewed the manuscript following co-author revision

RM reviewed the study design, cleaned the data and analyzed the data

NM is the study coordinator who reviewed the study implementation, data collection, coding and analysis, and reviewed/revised study drafts

ML reviewed the study design, reviewed the study data analysis and revised the final manuscript

JM reviewed the study design, study data and revised the final manuscript

LGB reviewed the study design, study data and revised the final manuscript

PG reviewed the study design, study data and revised the final manuscript

TC designed the study, reviewed the study data and revised the final manuscript

LM designed the study, reviewed the study data and revised the final manuscript

## Acknowledgments

We would like to thank our study participants, PrEP-PP study staff and the City of Cape Town Department of Health staff.

## Funding

This study was supported through grants from the National Institute of Mental Health (TC and LM; R01MH116771) and Fogarty International Center (DJD; K01TW011187).

## Notes

### Competing Interest Statement

The authors have declared no competing interest.

### Clinical Trial

NCT03826199

